# Comparative Value of Cognitive and Functional Assessments for Predicting 24-Month Progression from Mild Cognitive Impairment to Alzheimer’s Disease: An ADNI Cohort Study

**DOI:** 10.64898/2026.09.01.26360561

**Authors:** Sophie Choe

## Abstract

**Background:** Accurate prediction of progression from mild cognitive impairment (MCI) to Alzheimer’s disease (AD) is important for prognosis, patient management, and clinical trial enrollment.

**Objective:** Cognitive and functional assessments are routinely used in memory clinics, but their relative predictive value remains unclear. We sought to identify which assessments are most predictive of 24-month progression from MCI to AD.

**Methods:** We analyzed 2,430 participants with baseline MCI from the Alzheimer’s Disease Neuroimaging Initiative (ADNI) who were classified by 24-month progression to AD. Extreme Gradient Boosting (XGBoost) models were trained using repeated stratified 5-fold cross-validation with 10 repetitions. We compared demographic and genetic variables, global cognitive measures (MMSE, ADAS-Cog13, CDR-SB, MoCA), episodic memory, executive function, functional status, and Everyday Cognition (ECog) questionnaires.

**Results:** The baseline clinical model (age, sex, education, APOE ε4 status) achieved an area under the receiver operating characteristic curve (AUC) of 0.692. Episodic memory showed the highest predictive performance (AUC = 0.915), followed by the Functional Activities Questionnaire (AUC = 0.913).

Combining episodic memory, functional assessment, and executive function achieved the best performance (AUC = 0.943, sensitivity = 0.857, specificity = 0.889). Among individual memory measures, Logical Memory Delayed Recall achieved the highest standalone performance (AUC = 0.896), whereas RAVLT Learning provided minimal incremental value.

**Conclusions:** Episodic memory is the strongest predictor of 24-month progression from MCI to AD, with functional assessment providing substantial complementary value. Streamlined assessment batteries emphasizing episodic memory and functional status may improve efficient risk stratification in memory clinics and AD clinical trials.

## Background

Alzheimer’s disease (AD) is the leading cause of dementia worldwide and is characterized by a prolonged preclinical phase followed by progressive cognitive and functional decline [1]. Individuals diagnosed with mild cognitive impairment (MCI) represent a heterogeneous population: while some progress to AD within several years, others remain cognitively stable, revert to normal cognition, or develop alternative neurodegenerative disorders [2–5]. Identifying individuals at greatest risk of progression is therefore important for patient counseling, prognostic assessment, clinical trial enrollment, and timely implementation of disease-modifying therapies [6–8].

Current prediction of MCI progression draws on clinical evaluation, neuropsychological testing [9–11], neuroimaging [12], fluid biomarkers [13,14], and genetic information [15,16]. Although biomarkers such as amyloid PET [17,18], CSF analytes [14], and plasma phosphorylated tau [19,20] provide informative measures of AD pathology, their use may be limited by cost, invasiveness, accessibility, or the need for specialized infrastructure. Consequently, cognitive and functional assessments remain central components of clinical evaluation in memory clinics and research settings [9–11].

Commonly used instruments include global measures such as the MMSE [21], MoCA [22], ADAS-Cog [23], and CDR-SB [24], alongside domain-specific tests targeting episodic memory, executive function, and everyday functioning. Episodic memory tests such as the RAVLT [25] and Logical Memory [26] are especially relevant to prodromal AD, since episodic memory impairment is a prominent early feature of the disease [9]. Functional measures such as the FAQ [27] and patient-and informant-reported Everyday Cognition (ECog) questionnaires [28] provide complementary information and have shown prognostic value for subsequent clinical progression [29–32].

Prior statistical and machine learning studies, including our own, have shown that cognitive and functional assessments can provide strong predictive information for MCI-to-AD progression [33–38]. However, these studies have generally evaluated broad predictor sets rather than isolating individual assessments, leaving unclear which specific tests drive this predictive value and which may be redundant.

We therefore set out to systematically compare routinely used cognitive and functional assessments for predicting 24-month MCI-to-AD progression in 2,430 participants from the Alzheimer’s Disease Neuroimaging Initiative (ADNI) [39], first evaluating major assessment domains and then performing feature-ablation analyses to quantify the unique contribution of individual assessments.

## Methods

### Study Cohort

Data were obtained from the Alzheimer’s Disease Neuroimaging Initiative (ADNI) [39], a multicenter longitudinal study designed to identify clinical, imaging, genetic, and biochemical biomarkers for the early detection and tracking of Alzheimer’s disease (AD). Participants diagnosed with mild cognitive impairment (MCI) at baseline were included in this study. Individuals were classified as progressors if they converted to a clinical diagnosis of AD within 24 months of the baseline visit, and as non-progressors if they remained diagnosed with MCI throughout the same follow-up period.

Participants with missing outcome labels or insufficient follow-up to determine 24-month progression status were excluded. The final study cohort comprised 2,430 baseline MCI participants, including both progressors and non-progressors.

### Cognitive and Functional Assessments

Baseline cognitive and functional assessments were extracted from the ADNIMERGE dataset. To evaluate the relative contribution of different assessment domains, cognitive measures were grouped according to their primary clinical purpose, as described below.

#### Clinical Variables

The baseline clinical model included age, sex, years of education, and APOE ε4 carrier status.

#### Global Cognitive Measures

Global cognitive assessments included the Mini-Mental State Examination (MMSE), the 13-item Alzheimer’s Disease Assessment Scale–Cognitive Subscale (ADAS-Cog13), the Clinical Dementia Rating Sum of Boxes (CDR-SB), and the Montreal Cognitive Assessment (MoCA). Because MoCA was introduced after ADNI1, analyses involving MoCA were restricted to the subset of participants with available measurements.

#### Episodic Memory

The episodic memory domain included four measures: Rey Auditory Verbal Learning Test (RAVLT) Immediate Recall, RAVLT Learning, RAVLT Forgetting, and Logical Memory Delayed Recall (LDELTOTAL). These measures capture complementary aspects of verbal episodic memory, including immediate recall, learning efficiency, retention, and delayed recall.

#### Executive Function

Executive function was represented by Trail Making Test Part B completion time (TRABSCOR).

#### Functional Status

Functional performance was assessed using the Functional Activities Questionnaire (FAQ).

#### Everyday Cognition

Patient-and study partner-reported cognition were evaluated using the Everyday Cognition (ECog) questionnaire. Three ECog-based feature sets were examined: patient-reported ECog Total score, study partner-reported ECog Total score, and patient–study partner discrepancy scores. Because ECog was introduced after ADNI1, these analyses were also restricted to the subset of participants with available data.

### Experimental Design

To quantify the predictive value of individual assessment domains, multiple feature sets were evaluated independently. Primary experiments compared the following models: clinical variables alone; clinical + MMSE; clinical + ADAS-Cog13; clinical + CDR-SB; clinical + episodic memory; clinical + FAQ; clinical + executive function; clinical + episodic memory + FAQ; and clinical + episodic memory + FAQ + executive function. Secondary analyses evaluated MoCA-and ECog-based models using the corresponding data subsets.

### Memory Feature Contribution Analysis

Because episodic memory emerged as the strongest-performing individual assessment domain in this analysis, additional analyses examined the contribution of individual memory assessments. First, separate models were trained using each memory test individually, in combination with the baseline clinical variables (clinical + RAVLT Immediate Recall; clinical + RAVLT Learning; clinical + RAVLT Forgetting; clinical + LDELTOTAL). A combined memory model incorporating all four episodic memory measures was also evaluated.

Second, leave-one-feature-out ablation analyses were performed using the best-performing cognitive model (clinical + episodic memory + FAQ + executive function), with individual memory variables removed one at a time to quantify their unique contribution to predictive performance.

### Machine Learning Model

All experiments used Extreme Gradient Boosting (XGBoost), a tree-based ensemble algorithm that has demonstrated strong performance on structured clinical datasets. Class imbalance was addressed by computing the scale_pos_weight parameter within each training fold.

Categorical variables were one-hot encoded; continuous variables were imputed using the training-fold median, and missing categorical values were imputed using the most frequent category. All preprocessing steps were performed exclusively within training folds to prevent information leakage.

### Model Evaluation

Model performance was evaluated using repeated stratified five-fold cross-validation with ten repetitions (50 train–test splits total) [40]. The same cross-validation strategy was applied across all experiments to enable direct comparison between feature sets.

For each training fold, the optimal classification threshold was determined using Youden’s index [41] calculated from training-fold predictions and then applied to the corresponding test fold. Performance was summarized by averaging results across all cross-validation folds.

### Performance Metrics

The primary outcome measure was the area under the receiver operating characteristic curve (AUC) [42]. Secondary metrics included accuracy, balanced accuracy [43], sensitivity, specificity, precision, negative predictive value (NPV), F1-score, and Brier score [44]. Ninety-five percent confidence intervals for AUC were estimated by nonparametric bootstrap resampling of participants using the out-of-fold predictions [45].

### Statistical Comparison

The relative contribution of each cognitive domain was assessed by comparing cross-validated predictive performance across feature sets. For episodic memory analyses, both standalone model performance and the change in AUC following leave-one-feature-out ablation were examined to quantify the unique predictive value of individual memory assessments beyond the remaining cognitive measures.

## Results

### Study Cohort

The study included 2,430 participants with mild cognitive impairment (MCI) at baseline who had sufficient follow-up to determine 24-month progression status. All primary analyses used the full cohort. Analyses involving the Montreal Cognitive Assessment (MoCA) and Everyday Cognition (ECog) questionnaire were restricted to the subset of participants with available assessments, as these instruments were introduced after the ADNI1 phase.

Baseline demographic, clinical, and cognitive characteristics of the cohort, stratified by 24-month progression status, are presented in Table 1.

**Table 1.** Baseline characteristics of the study cohort, by 24-month progression status.

| Characteristic | Non-progressor<br>(n=1883) | Progressor<br>(n=547) | Overall (N=2430) | p |
| --- | --- | --- | --- | --- |
| N | 1883 | 547 | 2430 |  |
| Age, years, mean (SD) | 72.5 (7.2) | 74.3 (7.7) | 72.9 (7.4) | <0.001 |
| Female sex, n (%) | 927 (49.2%) | 230 (42.0%) | 1157 (47.6%) | 0.004 |
| Education, years, mean (SD) | 16.2 (2.7) | 15.5 (2.9) | 16.1 (2.7) | <0.001 |
| APOE ε4 carrier (≥1 allele), n (%) | 651 (38.8%) | 363 (68.1%) | 1014 (45.8%) | <0.001 |
| MMSE score, mean (SD) | 28.2 (2.0) | 24.5 (2.7) | 27.4 (2.7) | <0.001 |
| CDR-SB score, mean (SD) | 0.85 (1.18) | 3.58 (1.84) | 1.46 (1.77) | <0.001 |
| FAQ score, mean (SD) | 1.7 (3.7) | 10.9 (7.1) | 3.8 (6.0) | <0.001 |
| Logical Memory Delayed Recall, mean (SD) | 9.4 (4.9) | 2.1 (2.6) | 7.8 (5.4) | <0.001 |
| RAVLT Immediate Recall, mean (SD) | 40.3 (11.7) | 24.2 (7.5) | 36.7 (12.8) | <0.001 |

Values are mean (SD) for continuous variables and n (%) for categorical variables. p-values are from Welch's t-test (continuous variables) or chi-square test (categorical variables) comparing progressors and non-progressors. APOE4 status was missing for 217 participants (203 non-progressors, 14 progressors) and percentages are of participants with available genotype data. MMSE = Mini-Mental State Examination; CDR-SB = Clinical Dementia Rating Sum of Boxes; FAQ = Functional Activities Questionnaire; RAVLT = Rey Auditory Verbal Learning Test.

### Comparison of Cognitive Assessment Domains

The predictive performance of individual cognitive and functional assessment domains is summarized in Table 2. Corresponding results are displayed graphically in Figure 1, and receiver operating characteristic curves for each predictor set are shown in Figure 2.

**Figure 1.**
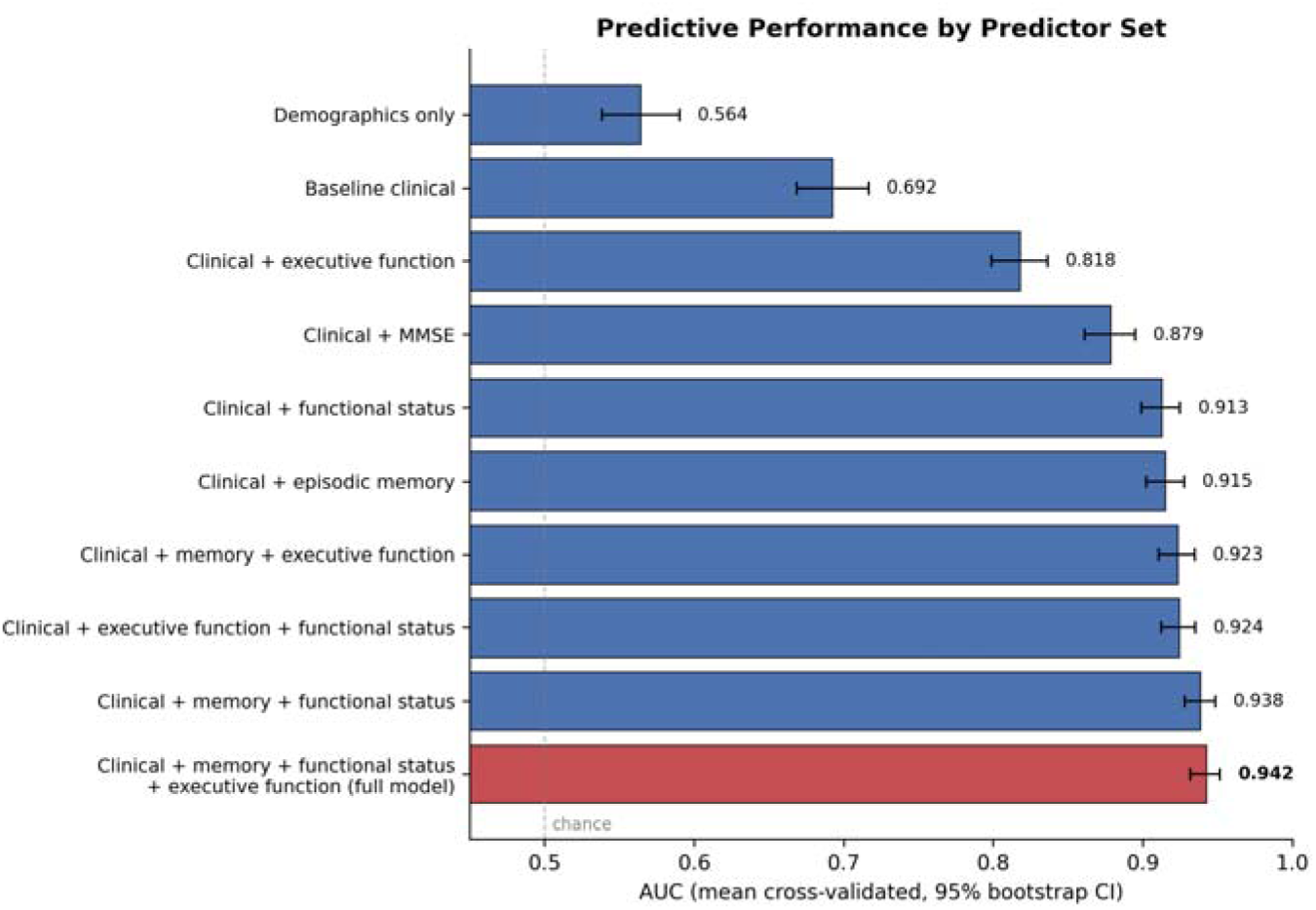
Predictive performance for 24-month progression from MCI to AD, by predictor set. Bars show mean cross-validated AUC (averaged across 50 repeated stratified five-fold cross-validation splits) for each predictor set, ordered ascending. Error bars show 95% CIs estimated by nonparametric bootstrap resampling of pooled out-of-fold predictions. The dashed line at 0.5 indicates chance-level discrimination. The full model (bottom, red) combines clinical variables, episodic memory, functional status, and executive function. MMSE = Mini-Mental State Examination.

**Figure 2.**
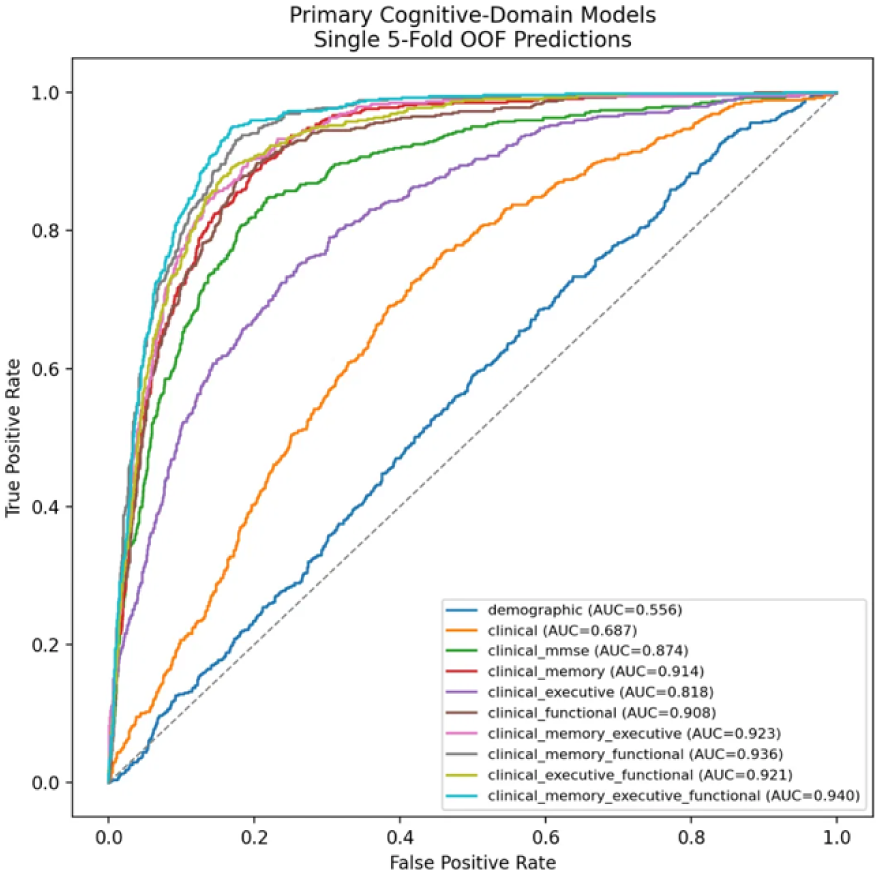
Receiver operating characteristic curves by predictor set. Receiver operating characteristic curves for each predictor set, derived from pooled out-of-fold predictions from a single representative five-fold cross-validation split. AUC values shown in the legend reflect this single split and differ slightly from the repeated cross-validation estimates (50 splits) reported in Table 2 and Figure 1. MMSE = Mini-Mental State Examination.

**Table 2.** Predictive performance for 24-month progression from MCI to AD, by predictor set.

| Model (predictor set) | Features (n) | AUC (95% CI) | Accuracy | Sensitivity | Specificity | Brier score |
| --- | --- | --- | --- | --- | --- | --- |
| Demographics only (age, sex, education) | 3 | 0.564 (0.530–0.582) | 0.557 | 0.521 | 0.567 | 0.241 |
| Baseline clinical (age, sex, education, APOE ε4) | 4 | 0.692 (0.664–0.712) | 0.649 | 0.642 | 0.650 | 0.213 |
| Clinical + executive function (TMT-B) | 5 | 0.818 (0.798–0.836) | 0.754 | 0.716 | 0.765 | 0.163 |
| Clinical + MMSE | 5 | 0.879 (0.856–0.890) | 0.804 | 0.825 | 0.798 | 0.132 |
| Clinical + functional status (FAQ) | 5 | 0.913 (0.895–0.921) | 0.843 | 0.833 | 0.846 | 0.113 |
| Clinical + episodic memory | 8 | 0.915 (0.901–0.926) | 0.841 | 0.842 | 0.841 | 0.112 |
| Clinical + episodic memory + executive function | 9 | 0.923 (0.910–0.934) | 0.857 | 0.841 | 0.861 | 0.104 |
| Clinical + executive function + functional status | 6 | 0.924 (0.909–0.932) | 0.859 | 0.837 | 0.866 | 0.103 |
| Clinical + episodic memory + functional status | 9 | 0.938 (0.926–0.946) | 0.869 | 0.848 | 0.875 | 0.095 |
| **Clinical + episodic memory + functional status + executive function (full model)** | **10** | **0.943 (0.929–0.949)** | **0.881** | **0.851** | **0.890** | **0.089** |

*Values are averaged across 50 repeated stratified five-fold cross-validation splits (10 repetitions × 5 folds); accuracy, sensitivity, and specificity use the Youden's-index threshold. 95% CIs for AUC were estimated by nonparametric bootstrap resampling of pooled out-of-fold predictions. The full model (bottom row, bolded) is shown for reference. AUC = area under the receiver operating characteristic curve; MMSE = Mini-Mental State Examination; TMT-B = Trail Making Test Part B; FAQ = Functional Activities Questionnaire.*

**Table 3.** Standalone performance and unique contribution of individual episodic memory measures.

| Memory measure | Standalone AUC<br>(clinical + measure alone) | Full-model AUC<br>with measure removed | ΔAUC<br>(unique contribution) |
| --- | --- | --- | --- |
| **Logical Memory Delayed Recall (LDELTOTAL)** | **0.896** | **0.936** | **0.006** |
| RAVLT Immediate Recall | 0.873 | 0.941 | 0.001 |
| RAVLT Forgetting | 0.720 | 0.941 | 0.001 |
| RAVLT Learning | 0.817 | 0.943 | -0.001 |

"Standalone AUC" reflects a model containing clinical variables plus that memory measure alone. "Full-model AUC with measure removed" reflects the leave-one-feature-out ablation: the full model (clinical + all four memory measures + functional status + executive function, AUC = 0.942) with the named measure excluded. $\Delta$ AUC is the full-model AUC (0.942) minus the ablated-model AUC, i.e., the unique predictive contribution of that measure within the full model; a negative value indicates the ablated model performed marginally better. For reference, the combined episodic-memory model (clinical + all four memory measures, without functional or executive predictors) achieved an AUC of 0.915. RAVLT = Rey Auditory Verbal Learning Test.

The baseline clinical model — age, sex, education, and APOE ε4 status — achieved an AUC of 0.692, indicating that demographic and genetic information alone provided only modest discrimination between progressors and non-progressors. Adding global cognitive assessments substantially improved performance: clinical variables combined with the Mini-Mental State Examination (MMSE) achieved an AUC of 0.879, while ADAS-Cog13 and CDR-SB further improved discrimination to AUCs of 0.900 and 0.911, respectively.

Among individual cognitive domains, episodic memory demonstrated the strongest predictive performance (AUC = 0.915). Functional assessment using the Functional Activities Questionnaire (FAQ) produced comparable discrimination (AUC = 0.913), whereas executive function, assessed by Trail Making Test Part B, achieved a lower AUC of 0.818.

Combining complementary domains further improved performance. Integrating episodic memory with functional assessment increased the AUC to 0.938, and the addition of executive function yielded the highest overall performance (AUC = 0.943, accuracy = 0.882, sensitivity = 0.857, specificity = 0.889, Brier score = 0.088). These findings indicate that episodic memory and functional assessment provide complementary prognostic information, while executive function contributes a modest additional improvement.

To isolate the contribution of genetic risk information, we additionally evaluated a demographics-only model (age, sex, education) without APOE ε4 status. This model performed poorly (AUC = 0.564), only modestly above chance, underscoring that demographic variables alone carry little predictive signal for MCI-to-AD progression. Adding APOE ε4 status to this same variable set — the baseline clinical model — increased AUC to 0.692, indicating that genetic risk information contributed more to prediction than age, sex, and education combined.

We also examined two additional two-domain combinations to further characterize executive function’s contribution. Adding executive function to the episodic memory model (clinical + memory + executive function) achieved an AUC of 0.923, a modest improvement over episodic memory alone (0.915) but smaller than the gain achieved by adding functional status (0.938). Similarly, combining executive function with functional status (clinical + executive function + functional status) achieved an AUC of 0.924, comparable to but not exceeding the memory + functional status combination (0.938). Together, these results suggest that executive function, while informative in isolation (AUC = 0.818), added limited incremental value once either episodic memory or functional status was already included in the model — consistent with its comparatively weaker standalone performance among the three domains.

### Performance of MoCA and ECog

Because MoCA and ECog were available only in participants enrolled during later ADNI phases, these analyses were performed on a reduced cohort and are not directly comparable to the primary analyses.

Combining MMSE with study partner-reported ECog achieved the strongest performance among these secondary analyses (AUC = 0.909), exceeding MMSE alone within the available subset. Study partner-reported ECog (AUC = 0.830) consistently outperformed patient self-reported ECog (AUC = 0.745), while ECog discrepancy scores showed intermediate performance (AUC = 0.798). MoCA alone achieved an AUC of 0.813, and combining MoCA with MMSE improved performance to an AUC of 0.895. Together, these findings suggest that informant-reported cognition provides greater prognostic value than patient self-report.

### Individual Episodic Memory Assessments

To determine the predictive value of individual memory assessments, separate models were constructed using each episodic memory measure in combination with the baseline clinical variables. Logical Memory Delayed Recall (LDELTOTAL) achieved the highest standalone performance among individual memory assessments (AUC = 0.896), followed by RAVLT Immediate Recall (AUC = 0.873) and RAVLT Learning (AUC = 0.817). RAVLT Forgetting showed substantially lower predictive performance (AUC = 0.720). Combining all four episodic memory measures increased the AUC to 0.915, indicating that the individual memory tests provide complementary information when evaluated together.

### Contribution of Individual Memory Assessments

To quantify the unique contribution of each memory assessment, leave-one-feature-out ablation analyses were performed using the best-performing model (clinical variables, episodic memory, FAQ, and executive function; full-model AUC = 0.943).

Removing Logical Memory Delayed Recall produced the largest reduction in performance, decreasing the AUC to 0.936. Removing RAVLT Immediate Recall or RAVLT Forgetting resulted in only modest decreases (AUC = 0.941 for both), and removing RAVLT Learning had no meaningful effect (AUC = 0.943), suggesting that its predictive information was largely redundant with the remaining memory assessments. These findings indicate that Logical Memory Delayed Recall provides the greatest unique contribution among the evaluated episodic memory measures, whereas RAVLT Learning contributes little additional prognostic information beyond the other memory tests.

## Discussion

In this study, we systematically compared the predictive value of routinely administered cognitive and functional assessments for forecasting progression from mild cognitive impairment (MCI) to Alzheimer’s disease (AD) within 24 months. By evaluating individual cognitive domains, combinations of assessments, and the contribution of specific episodic memory tests, we identified assessment strategies that maximize predictive performance while clarifying the relative importance of commonly used neuropsychological measures.

The baseline clinical model — age, sex, education, and APOE ε4 status — demonstrated only modest predictive performance. Although these variables are well-established risk factors for AD, they primarily reflect long-term susceptibility rather than imminent clinical progression. This finding underscores the importance of direct cognitive and functional assessment when estimating short-term conversion risk among individuals who already meet criteria for MCI.

Among the individual cognitive domains examined, episodic memory demonstrated the strongest predictive performance. This observation is consistent with the neuropathological progression of AD, in which early degeneration of medial temporal lobe structures produces prominent episodic memory impairment before more widespread cognitive decline becomes apparent. The superior performance of the memory model relative to global cognitive screening instruments such as MMSE suggests that targeted episodic memory assessment captures disease-specific information that is diluted in broader screening measures.

Functional assessment using the FAQ achieved predictive performance comparable to the episodic memory model and substantially improved performance when combined with memory measures. Functional decline reflects the real-world consequences of cognitive impairment and may capture subtle deficits not fully reflected in formal neuropsychological testing. The complementary improvement observed when memory and functional assessments were combined suggests that these domains provide distinct but synergistic prognostic information.

Executive function, represented by Trail Making Test Part B, showed more modest predictive performance independently but provided incremental benefit when added to the combined memory and functional model. This suggests that executive dysfunction contributes additional prognostic information beyond episodic memory, though its independent contribution is smaller than that of memory or functional status.

Among the global cognitive assessments, CDR-SB and ADAS-Cog13 outperformed MMSE. Although MMSE remains one of the most widely used cognitive screening instruments in clinical practice because of its simplicity and familiarity, it was less predictive than more comprehensive assessments of cognitive and functional impairment — consistent with prior reports of limited MMSE sensitivity for detecting subtle cognitive changes in prodromal AD.

Secondary analyses of MoCA and ECog should be interpreted cautiously, as these assessments were available only in later ADNI phases. Nevertheless, study partner-reported ECog consistently outperformed patient self-report, supporting prior observations that informants often provide more reliable assessments of cognitive decline than affected individuals, who may have reduced awareness of their own deficits in early AD.

A major contribution of this study is the detailed evaluation of individual episodic memory assessments. Logical Memory Delayed Recall demonstrated both the strongest standalone predictive performance and the largest unique contribution within the combined cognitive model, whereas RAVLT Learning contributed little additional predictive information after accounting for the remaining memory measures. These findings suggest that delayed story recall captures aspects of episodic memory particularly informative for identifying individuals at greatest risk of progression, and indicate that some components of traditional memory batteries may provide overlapping information — creating opportunities to simplify cognitive assessment without substantially compromising predictive accuracy.

From a clinical perspective, our findings support a hierarchical approach to cognitive assessment. When assessment time is limited, episodic memory testing should be prioritized, as it provides the greatest individual predictive value. Incorporating a brief functional assessment such as the FAQ substantially improves prediction, and adding executive function testing provides a modest further benefit. This progression suggests that a streamlined battery emphasizing memory and functional status may achieve predictive performance comparable to considerably larger neuropsychological batteries, potentially improving feasibility in routine clinical practice and resource-limited settings.

Strengths of this study include the use of a large, well-characterized multicenter cohort; systematic comparison of multiple cognitive domains within a consistent machine learning framework; repeated stratified cross-validation to improve the robustness of performance estimates; and feature ablation analyses quantifying the unique contribution of individual memory assessments. Rather than focusing solely on maximizing predictive accuracy, this study provides clinically interpretable evidence regarding the relative value of commonly administered cognitive assessments.

Several limitations should be acknowledged. First, all analyses used the ADNI cohort, which may not fully represent patients encountered in routine clinical practice, as participants are generally highly educated, predominantly recruited from research centers, and undergo standardized evaluations; external validation in more diverse, independent cohorts is needed [46]. Second, MoCA and ECog were introduced after the ADNI1 phase, resulting in smaller analytic samples for these analyses and limiting direct comparison with experiments performed on the full cohort. Third, although repeated cross-validation provides internal validation [40], prospective validation in real-world clinical settings is needed before these findings can inform routine patient care. Finally, this study focused exclusively on cognitive and functional assessments; future work should examine how optimized cognitive batteries can be integrated with structural MRI, plasma biomarkers, cerebrospinal fluid biomarkers, and positron emission tomography to further improve individualized risk prediction [12–20].

## Conclusions

In this study, we systematically evaluated the predictive value of routinely administered cognitive and functional assessments for forecasting progression from mild cognitive impairment (MCI) to Alzheimer’s disease (AD) within 24 months. Episodic memory, particularly Logical Memory Delayed Recall, emerged as the single strongest predictor of 24-month MCI-to-AD progression, with functional assessment providing substantial complementary value; combining the two — with a modest addition from executive function — achieved the best overall performance. These findings support prioritizing episodic memory and functional evaluation over broader screening batteries in memory clinics and AD trials, pending prospective validation in more diverse cohorts.

## Data Availability

Data Availability Statement: The datasets analyzed in the present study are available online from the Alzheimer's Disease Neuroimaging Initiative (ADNI) to qualified investigators upon application and approval: https://adni.loni.usc.edu

## Acknowledgments

Data collection and sharing for this project were funded by the Alzheimer’s Disease Neuroimaging Initiative (ADNI) (National Institutes of Health Grant U01 AG024904) and the Department of Defense ADNI (award number W81XWH-12-2-0012). ADNI is funded by the National Institute on Aging, the National Institute of Biomedical Imaging and Bioengineering, and through generous contributions from numerous public and private partners. The grantee organization is the Northern California Institute for Research and Education, and the study is coordinated by the Alzheimer’s Therapeutic Research Institute at the University of Southern California. ADNI data are disseminated by the Laboratory for Neuro Imaging at the University of Southern California.

The author thanks the investigators and participants of the Alzheimer’s Disease Neuroimaging Initiative for making this research possible. The ADNI investigators contributed to the design and implementation of ADNI and/or provided data but did not participate in the analysis or writing of this report. A complete listing of ADNI investigators is available at: https://adni.loni.usc.edu/wp-content/uploads/how_to_apply/ADNI_Acknowledgement_List.pdf.

## Author Contributions

Sophie Choe conceived and designed the study, curated the data, developed the methodology, performed the analyses, interpreted the results, created the figures and tables, and wrote, reviewed, and approved the final manuscript.

## Statements and Declarations Ethical considerations

This study is a secondary analysis of de-identified data obtained from the Alzheimer’s Disease Neuroimaging Initiative (ADNI). The ADNI study was approved by the Institutional Review Board at each participating institution, and all participants provided written informed consent. The present secondary analysis of de-identified publicly available data did not require additional institutional ethics approval.

## Consent to participate

Written informed consent was obtained from all participants by the Alzheimer’s Disease Neuroimaging Initiative (ADNI) at the participating study sites.

## Consent for publication

Not applicable.

## Data availability

The datasets analyzed during the current study are available from the Alzheimer’s Disease Neuroimaging Initiative (ADNI) to qualified investigators upon application and approval. Information regarding data access is available at https://adni.loni.usc.edu.

The code used to perform the analyses reported in this study is available from the corresponding author upon reasonable request.

